# Rethinking Prenatal Folate Assessment: Maternal Folate Receptor Autoantibodies and Child Neurodevelopment

**DOI:** 10.64898/2026.01.15.26344170

**Authors:** Qingyang Li, Wenyi Xu, Junjie Ao, Luan Chen, Cong Huai, Qianlong Zhang, Qiang Luo, Fei Li

## Abstract

Folic acid supplementation during pregnancy is established for preventing neural tube defects, yet its broader neurodevelopmental benefits remain inconsistently observed. Folate receptor autoantibodies (FRAA) may disrupt folate transport, but population evidence is lacking. In a prospective cohort of 3,006 mother-child pairs from the Shanghai Birth Cohort, we precisely quantified maternal red blood cell 5-methyltetrahydrofolate (active folate) and FRAA levels in early pregnancy. We found that higher maternal folate was associated with better offspring cognitive performance, especially in executive function. This benefit was attenuated by elevated maternal FRAA (≥70th percentile). Notably, the attenuating effect of FRAA was specifically amplified among mothers with higher milk intake. Our findings highlight the importance of considering maternal immune status in prenatal folate assessment and support personalized nutritional strategies that consider immune context to optimize offspring neurodevelopment.

## Introduction

Folic acid supplementation is a cornerstone of global prenatal care, with the World Health Organization recommending 400 µg/day to prevent neural tube defects(1). Despite widespread implementation, the broader neurodevelopmental benefits of prenatal folic acid, such as cognition and neurodevelopment, remain inconsistent(2-4), indicating that folate’s protective efficacy may depend on additional factors. Emerging evidence implicates that folate receptor autoantibodies (FRAA) can disrupt folate transport to the fetal brain in susceptible individuals(5), leading to a state of functional folate insufficiency in the developing fetal brain despite normal maternal serum levels. This reveals a critical limitation: normal serum folate concentrations do not guarantee functional folate adequacy within the fetal brain. Consequently, advancing our understanding of this mechanism is essential to inform and guide more effective global prenatal folate policies.

In literature, elevated FRAA levels in mothers have been associated with neural tube defects in their children(6). Meanwhile, high prevalence of FRAA was also reported in children with neurodevelopmental disorders (e.g., cerebral folate deficiency and autism spectrum disorder)(7, 8). These FRAAs can block folate transport across the placenta and the blood-brain barrier to the fetal brain(9). Most relevant to pregnancy, molecular mimicry has been hypothesized as a mechanism for FRAA production, as molecular mimicry triggered by soluble folate-binding protein (FBP) in milk (bovine) facilitates the production of FRAAs that cross-react with FRs(10, 11). This is supported by a population study in Spain shown that FRAA is present in men and women (n=787), and notably, high milk consumers among them have an increased risk of FRAA production(12).

A primary obstacle in elucidating this complex interaction has been the lack of not only FRAA data in large-scale, prospective cohorts but also a precise biomarker for functional folate status. Conventional clinical kits measure "total folate," a conflated metric that does not reflect the bioactive folate fraction, potentially leading to biased associations in neurodevelopmental outcomes(13). A strength of our study is that, within the Shanghai Birth Cohort (SBC), we utilized our self-developed and patented mass spectrometry method to precisely quantify 5-methyltetrahydrofolate (5-MTHF), the predominant active form of folate, in maternal red blood cells (RBC). Mass spectrometry, considered the gold standard for metabolite detection, allows for the specific separation of 5-MTHF from other folate metabolites(13). Thus, while direct assessment of functional folate status in the fetal brain remains unfeasible, the precise quantification of 5-MTHF in maternal RBCs provides a superior proxy for functional folate availability at the maternal-fetal interface, overcoming key limitations of conventional serum total folate assays. This enables our study to delineate the association between active folate and offspring neurodevelopment in a large population, thereby providing reliable evidence for clarifying the role of FRAA. Thus, the SBC(14), with its 3,006 mother-child pairs and the novel biomarker data, serves as an ideal platform to investigate these compelling questions.

In this study, we tested the hypothesis that maternal FRAA moderates the beneficial effects of maternal folate levels on child neurodevelopment. In the SBC, we measured maternal RBC folate and FRAA levels in early pregnancy and systematically evaluated offspring neurodevelopment at birth and at 2, 4, and 7 years of age. To optimize prenatal folate management with attention to the potential impact of FRAA, we asked the following questions: 1) What are the most significant benefits of maternal folate supplementation among broader neurodevelopment outcomes? 2) Can the maternal FRAA moderate these associations? 3) Whether nutrition intake, such as milk intake, can interact with these FRAA moderation effects?

## Materials and Methods

### Study Population

The SBC is a prospective study established between 2013 and 2016, comprising a mother-child cohort recruited from six hospitals in Shanghai(14). Eligible participants met the following criteria: (1) women aged ≥20 years; (2) at least one partner registered as a Shanghai resident; (3) planned to seek prenatal care and deliver at SBC-participating hospitals; (4) intended to remain in the catchment area for at least two years; and (5) provided informed consent for longitudinal follow-up. Written informed consent was obtained from all participants, and the study protocol was approved by the Medical Ethics Committee of Shanghai Xinhua Hospital, Shanghai Jiao Tong University School of Medicine (Approval Number: XHEC-C-2013-001).

A total of 4,127 participant couples were initially enrolled in the SBC. After excluding 303 couples lost to follow-up, 125 miscarriages, and 7 stillbirths, 3,692 mother-newborn pairs with live births were identified. From this group, the final analytical sample for the primary analysis comprised 3,006 mother-newborn pairs who had complete data on both early-pregnancy RBC folate and FRAA. Subsequent analysis on a specific moderator (i.e., milk intake) was conducted on a subset of this primary sample with available data, with the exact sample size specified in Supplementary Table 1-2 and Supplementary Fig. 1.

### Measures

#### Assessment of RBC Folate

5-MTHF, the predominant and biologically active form of folate in humans, is stable in RBC, making it an ideal biomarker that reflects long-term folate stores(13, 15). Levels of RBC 5-MTHF in both maternal (early pregnancy) and cord blood samples were quantified using liquid chromatography-tandem mass spectrometry (LC-MS/MS) with a Complete Folate-function Test Kit (Vito Diagnostics, Hangzhou, China).

Briefly, after centrifugation of venous blood, the packed RBC was subsequently washed twice with physiological saline to obtain purified samples for analysis. A 60 μL aliquot of the RBC sample, calibrator, or quality control was mixed with 60 μL of a stable isotope-labeled internal standard solution and 180 μL of sample extraction solution. The mixture was incubated at 37°C for 60 minutes in the dark. Subsequently, 300 μL of deproteinizing solution was added, and the mixture was centrifuged at 2000 × g for 20 minutes. Then, 8 μL of the supernatant was injected into the LC-MS/MS system for analysis. Chromatographic separation was performed on a reversed-phase C18 column with a mobile phase of 0.1% formic acid in water and methanol, using a gradient elution at a flow rate of 0.5 mL/min. Mass spectrometric detection was carried out in positive electrospray ionization mode with multiple reaction monitoring (MRM). The MRM transitions were m/z 460.2 → 313.0 for 5-MTHF and 465.2 → 313.0 for the internal standard. Data acquisition and quantification were processed using Analyst MD 1.6.3 and MultiQuant MD 3.0.3 software, respectively.

To ensure the accuracy and reproducibility of the method, each analytical batch included calibration curves and quality controls (QCs). The method demonstrated good linearity (r > 0.99) over the quantifiable range of 15.6 to 2000 ng/mL, with a limit of detection (LOD) of 7.81 ng/mL for 5-MTHF. The accuracy was assessed using QCs at low (400 ng/mL) and high (1700 ng/mL) concentrations, yielding recoveries between 80% and 120%. The precision, expressed as the coefficient of variation (CV) of QCs, was below 20%.

#### Measurement of FRAA

FRAA levels in maternal serum samples were measured using a Human FRAA Enzyme-Linked Immunosorbent Assay (ELISA) Kit (Jianglai Biotechnology, Shanghai, China), according to the manufacturer’s instructions. Briefly, 50 μL of standards and test serum samples were added to the sample wells, followed by 100 μL of horseradish peroxidase-conjugated detection antibody. The plate was sealed and incubated at 37°C for 60 minutes. After incubation, the plate was washed five times. Subsequently, 50 μL each of Substrate A and Substrate B were added to each well, and the plate was incubated at 37°C for 15 minutes in the dark. The enzymatic reaction was terminated by adding 50 μL of Stop Solution, which changed the color from blue to yellow. The optical density at 450 nm was measured within 15 min. A standard curve was generated to calculate the serum FRAA concentration. The LOD for the FRAA assay was 1.0 ng/mL. The intra- and inter-assay CV of the kit were <9% and <11%, respectively.

#### Assessment of Covariates and Potential Modifiers

Trained interviewers conducted in-person interviews using standardized questionnaires to collect demographic information, including maternal sociodemographic characteristics, educational attainment, and occupation during early pregnancy. Among the potential modifiers considered, we focused on maternal milk intake, which was assessed through a self-report dietary habits survey. Intake frequency was coded as 0 (never), 1 (<1 time per week), 2 (1-3 times per week), 3 (4-7 times per week), and 4 (≥2 times per day). This assessment captured conventional milk products rather than specialized hydrolyzed formulas.

#### Child Neurodevelopmental Outcomes

Data on parity, pregnancy complications, and birth outcomes (birth weight, length, and head circumference) were abstracted from medical records. All neonatal measurements were obtained immediately after delivery. Birth weight and length were measured using an electronic infant scale (SECA 418, Seca GmbH & Co. KG, Germany), and head circumference was measured with a flexible tape from the frontal bone to the occipital protuberance.

Children were followed at 2, 4, and 7 years of age. At these visits, standardized developmental and behavioral measures were administered. While existing literature has established a link between maternal folate and general cognitive development, the evidence remains inconsistent, in part due to reliance on broad composite scores and lack of examination of specific domains(16). Our analysis using detailed neuropsychological tools was designed to address the gap in understanding which specific cognitive domains (*e.g.*, executive functions, memory, reasoning) are most susceptible to maternal folate status. Developmental and cognitive functioning was assessed through examiner-administered tests, including the Bayley Scales of Infant Development (Bayley)(17), the Wechsler Intelligence Scale for Children (WISC)(18), and the Behavior Rating Inventory of Executive Function (BRIEF)(19). Behavioral and socioemotional functioning was evaluated using validated parent report questionnaires, including the Strengths and Difficulties Questionnaire (SDQ)(20) and the Social Responsiveness Scale (SRS)(21).

Functional near-infrared spectroscopy (fNIRS) data were acquired at the 4-year and 8-year follow-up visits using a NirScan-900D system (Danyang Huichuang Medical Equipment Co., Ltd., Jiangsu, China). Data were collected during both a resting-state period and a Go/No-Go task. For each condition, concentration changes of oxygenated hemoglobin were calculated from the raw optical intensity data using the modified Beer-Lambert law. Detailed preprocessing steps, including artifact correction and analytical procedures, are described in the Supplementary Methods.

### Statistical Analysis

All analyses were performed using R software (version 4.3.3). For all continuous variables, values exceeding four standard deviations from the mean were considered outliers and set to missing (NA) prior to analysis. False discovery rate (FDR) correction was applied for multiple testing across neurodevelopmental phenotypes where indicated.

#### Association Analysis

We first conducted correlation analysis to evaluate the associations between maternal RBC folate (continuous) and child neurodevelopmental phenotypes. All models were adjusted for offspring sex and the interval between RBC folate measurement and sample collection.

#### Phenotype Set Enrichment Analysis

To identify which broad neurodevelopmental domains were most strongly associated with maternal RBC folate levels, we performed a phenotype set enrichment analysis. This approach, inspired by the logic of Gene Set Enrichment Analysis (GSEA)(22), groups individual measurement variables (*e.g.*, specific cognitive test scores, behavioral questionnaire subscores, growth parameters, and fNIRS metrics) into predefined phenotypic sets representing core domains: Cognition, Behavior, Growth, and Brain (fNIRS). First, each individual variable was ranked based on the strength and direction of its association with RBC folate, using a composite statistic (*e.g.*, -log_10_(p-value) * sign(beta coefficient)). Then, for each phenotypic set, we tested whether its member variables were non-randomly clustered toward the top (or bottom) of this ranked list, indicating a domain-wide association with folate. This strategy shifts the inference from numerous individual tests to a few domain-level tests, reducing the burden of multiple comparisons and enhancing the interpretability of findings.

#### Definition of FRAA Grouping (Exploratory Threshold Scanning)

Given the lack of a consensus clinical cutoff for FRAA in this context, we conducted an exploratory analysis to identify a level indicative of potential biological interference. We systematically tested FRAA percentiles from the 50th to the 90th as potential cutoffs, with multiple testing corrected using the FDR method. The threshold that yielded the most statistically significant interaction effect with RBC folate on the primary cognitive outcome was selected for subsequent moderation analyses. We emphasize that this analysis is hypothesis-generating, and the identified threshold requires validation in independent cohorts.

#### Moderation Analysis

To examine the potential enhancement of antibody interference by maternal milk intake, we adopted a stratified analysis approach. Milk intake frequencies (scores 0-4) were categorized into low intake (scores 0-2) and high intake (scores 3-4) groups based on established dietary assessment practices. Within each milk stratum, we then fitted multiple linear regression models to test the interaction between RBC folate and FRAA grouping on neurodevelopmental outcomes, adjusting for child sex, the interval between RBC folate measurement and sample collection. This approach avoids the statistical complexities of three-way interactions while providing robust evidence for effect modification.

#### Sensitivity Analysis

To assess the robustness of our primary findings, we conducted sensitivity analyses by further adjusting the main association and moderation models for potential confounding factors, including maternal age at delivery, educational attainment, and fetal birth type (singleton vs. multiple). The effect estimates for the associations between RBC folate and neurodevelopmental outcomes, as well as the interaction effects involving FRAA, remained consistent and statistically significant after these adjustments, confirming the stability of our conclusions.

#### Genetic Analyses Using GWAS and Mendelian Randomization

A two-stage genetic analysis was performed to investigate the genetic underpinnings of FRAA and a possible causal link with milk intake. First, a genome-wide association study (GWAS) was conducted among 1,635 Han Chinese female SBC participants to identify genetic variants associated with FRAA concentrations. Genotyping was performed on the Illumina GSA-24v3-0-EA array, followed by phasing, imputation against the 1000 Genomes Phase 3 reference panel, and rigorous quality control. A linear regression model, adjusted for age and genetic principal components, was used for association testing. Genome-wide significance was defined as p < 5 × 10⁻⁸.

Subsequently, a two-sample Mendelian Randomization (MR) analysis was employed to assess the causal effect of milk intake on FRAA levels. Genetic instruments for milk intake were obtained from the UK Biobank, and the inverse-variance weighted (IVW) method served as the primary analysis. Robustness was evaluated using MR-Egger and weighted median estimators, and sensitivity analyses were conducted to assess pleiotropy and heterogeneity. Full methodological details for genotyping, quality control, and MR procedures are provided in the Supplementary Methods.

## Results

### Descriptive Statistics

Among the 3,006 enrolled mother–newborn pairs in the analytical cohort, the mean maternal age at delivery was 28.98 years (*SD* = 3.64). A detailed summary of maternal characteristics, child outcomes, and biochemical measures is presented in Table 1 and Supplementary Table 3.

**Table 1.**
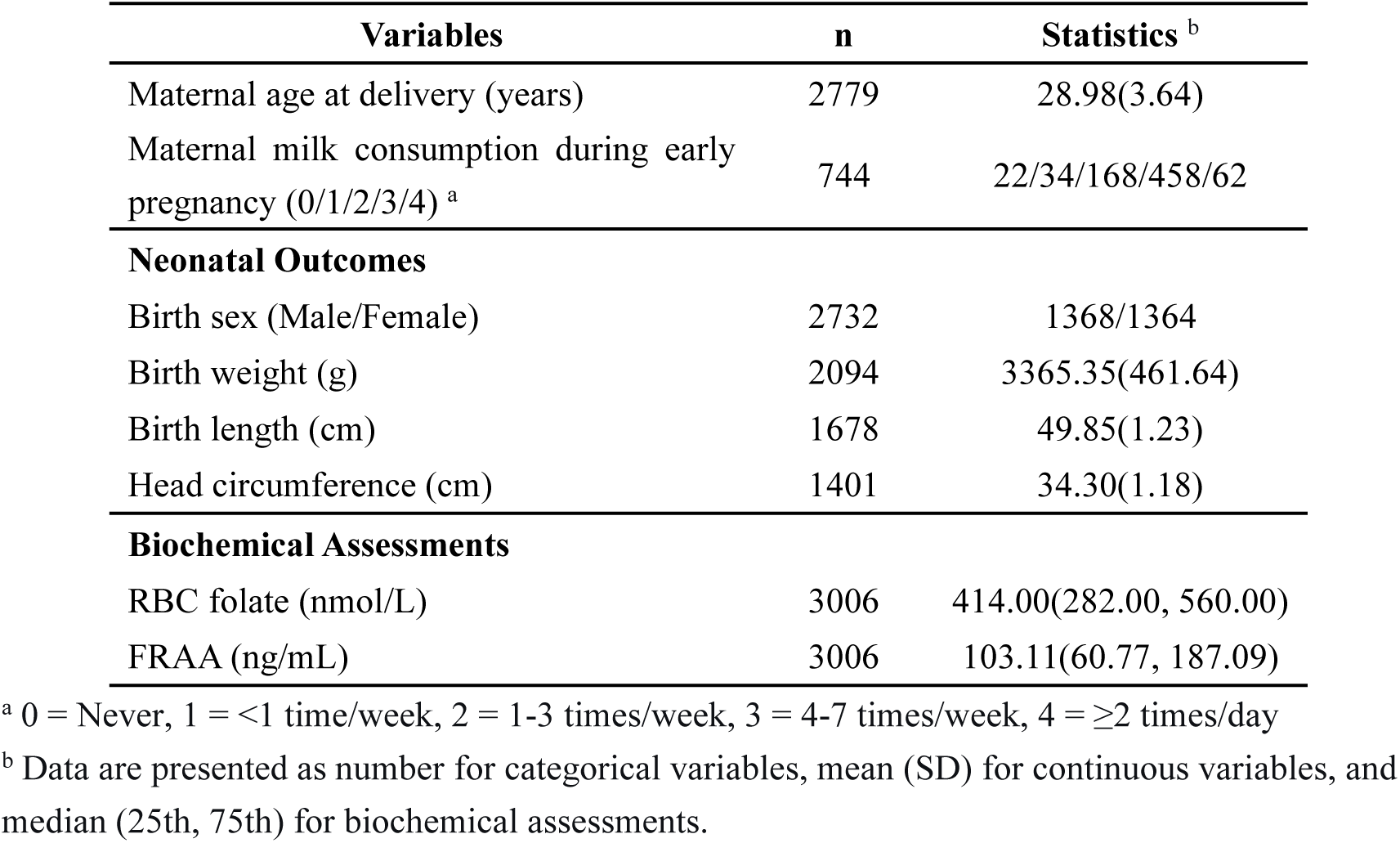
Presents detailed descriptive statistics for all key maternal and child measures.

| Variables | n | Statistics <sup>b</sup> |
| --- | --- | --- |
| Maternal age at delivery (years) | 2779 | 28.98(3.64) |
| Maternal milk consumption during early pregnancy (0/1/2/3/4) <sup>a</sup> | 744 | 22/34/168/458/62 |
| **Neonatal Outcomes** |  |  |
| Birth sex (Male/Female) | 2732 | 1368/1364 |
| Birth weight (g) | 2094 | 3365.35(461.64) |
| Birth length (cm) | 1678 | 49.85(1.23) |
| Head circumference (cm) | 1401 | 34.30(1.18) |
| **Biochemical Assessments** |  |  |
| RBC folate (nmol/L) | 3006 | 414.00(282.00, 560.00) |
| FRAA (ng/mL) | 3006 | 103.11(60.77, 187.09) |

<sup>a</sup> 0 = Never, 1 = <1 time/week, 2 = 1-3 times/week, 3 = 4-7 times/week, 4 = ≥2 times/day
<sup>b</sup> Data are presented as number for categorical variables, mean (SD) for continuous variables, and median (25th, 75th) for biochemical assessments.

### Associations Between Maternal RBC Folate Levels and Offspring Neurodevelopmental Outcomes

We found that higher maternal RBC folate concentrations were associated with both higher offspring cognitive scores and improved growth outcomes (*p* < 0.05, Fig. 1A, Supplementary Table 4). Across the four neurodevelopmental domains, folate associations were significantly enriched in the cognitive domain (Fig. 1B). This was further supported by Bayley scores at age 2, which confirmed associations with cognitive (*β* = 0.061, 95% CI: [0.017,0.107], *p* = 0.030, Fig. 1C) and language abilities (*β* = 0.057, 95% CI: [0.013,0.102], *p* = 0.030, Fig. 1C), but not with behavioral outcomes (including motor, social-emotional, and adaptive behavior). To further specify which cognitive dimensions were involved, we analyzed WISC scores at age 4 and found that folate levels were linked to fluid reasoning (*β* = 0.060, 95% CI: [0.007,0.112], *p* = 0.027, Fig. 1D), which is a component of executive function. Building on this, we examined more detailed aspects of executive function using the BRIEF at age 7. The results showed that maternal RBC folate was associated with the Metacognition Index (*β* = 0.064, 95% CI: [0.008,0.116], *p* = 0.024, Fig. 1E), as well as its constituent components, including task initiation, planning, organization of materials, and self-monitoring (Supplementary Table 4).

**Fig. 1.**
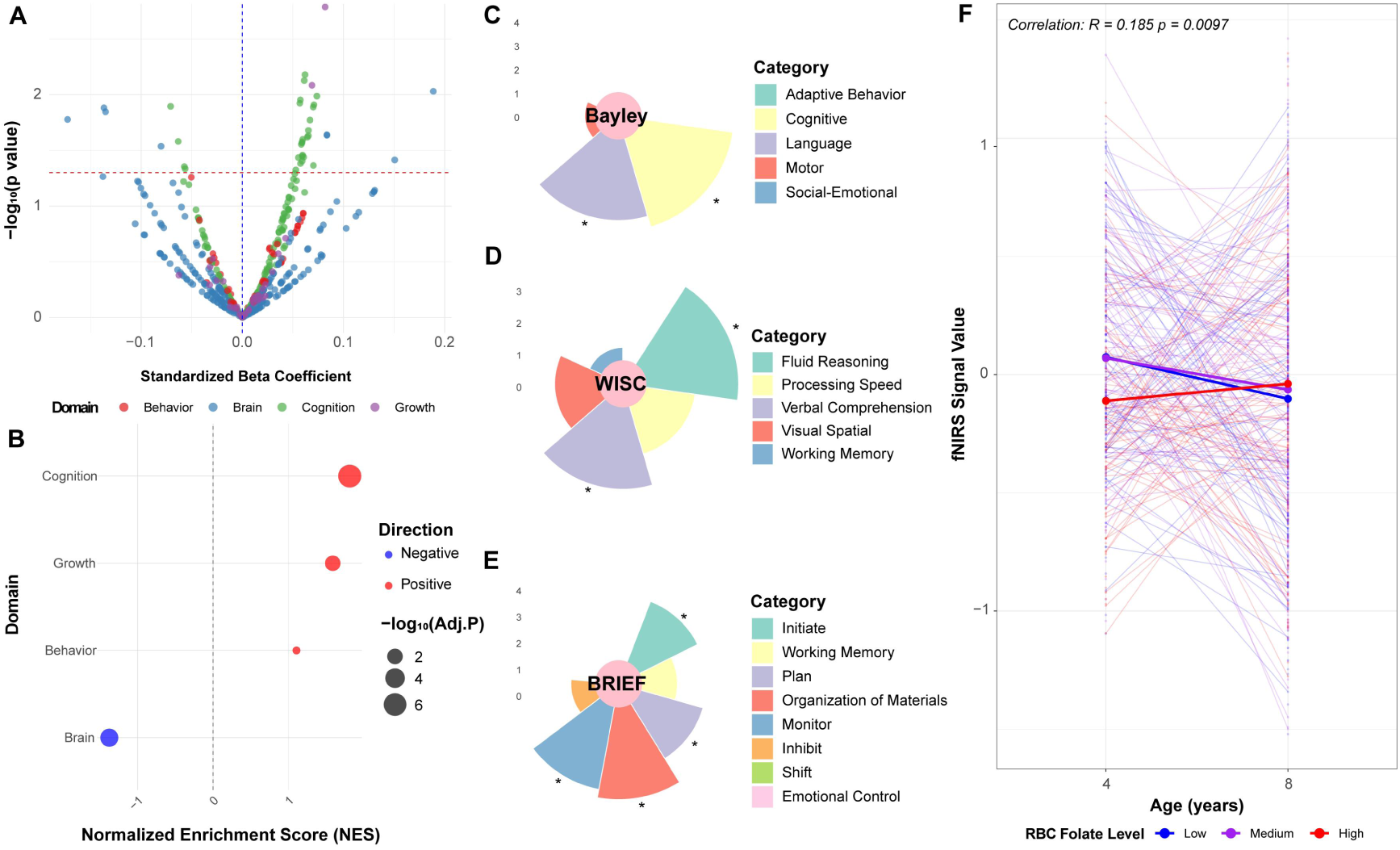
Associations between maternal early-pregnancy RBC folate and offspring neurodevelopmental and growth outcomes. (**A**) Volcano plot displaying the strength of association (-log₁₀ p-value) between maternal RBC folate and a spectrum of offspring neurodevelopmental and growth phenotypes. (**B**) Gene Set Enrichment Analysis (GSEA) results demonstrating that maternal RBC folate is most strongly enriched for associations in the cognitive domain, surpassing other phenotypic domains such as behavior and growth. (**C**-**E**) Radial bar plots where each sector represents a specific cognitive test metric. The radial height of the sector corresponds to the beta coefficient (quantitative measure of association strength), and the color denotes the qualitative cognitive domain to which the metric belongs (*e.g.*, Executive Function, Memory, Reasoning). An asterisk at the tail of a sector indicates that the association is statistically significant (p < 0.05): (**C**) Bayley Scales of Infant and Toddler Development (Bayley) at age 2. (**D**) Wechsler Intelligence Scale for Children (WISC) indices at age 4. (**E**) Behavior Rating Inventory of Executive Function (BRIEF) indices at age 7. (**F**) Developmental trajectories of task-evoked fNIRS activation in the left middle frontal gyrus (channel 7) from age 4 to 8 years, stratified by maternal folate levels.

Supporting these cognitive associations, fNIRS data revealed that children of mothers with higher folate levels showed a steeper developmental increase in task-related hemodynamic response in the left middle frontal gyrus (channel 7) from ages 4 to 8 years, compared to those with medium or low folate levels (*β* = 0.160, 95% CI: [0.040,0.281], *p* = 0.014, Fig. 1F, Supplementary Table 4, Supplementary Fig. 2-3). Resting-state analyses further suggested additional associations between folate and connectivity within the default mode, sensorimotor and visual networks (positive correlations at age 4, mixed positive and negative at age 8), though these did not survive strict correction for multiple comparisons (Supplementary Table 4).

### Folate Receptor Autoantibodies Moderate the Neurodevelopmental Benefits of Maternal Folate

Threshold scanning identified the 70th percentile of FRAA levels (163.87 ng/mL) as the cutoff yielding the most significant interaction with RBC folate on cognitive outcomes. Using this threshold to dichotomize participants, we found that the interaction term between RBC folate and the FRAA group was statistically significant and negative for both Bayley cognitive scores (*β* = -0.147, *p* = 0.027; Fig. 2A) and language scores (*β* = -0.173, *p* = 0.008; Fig. 2B).

**Fig. 2.**
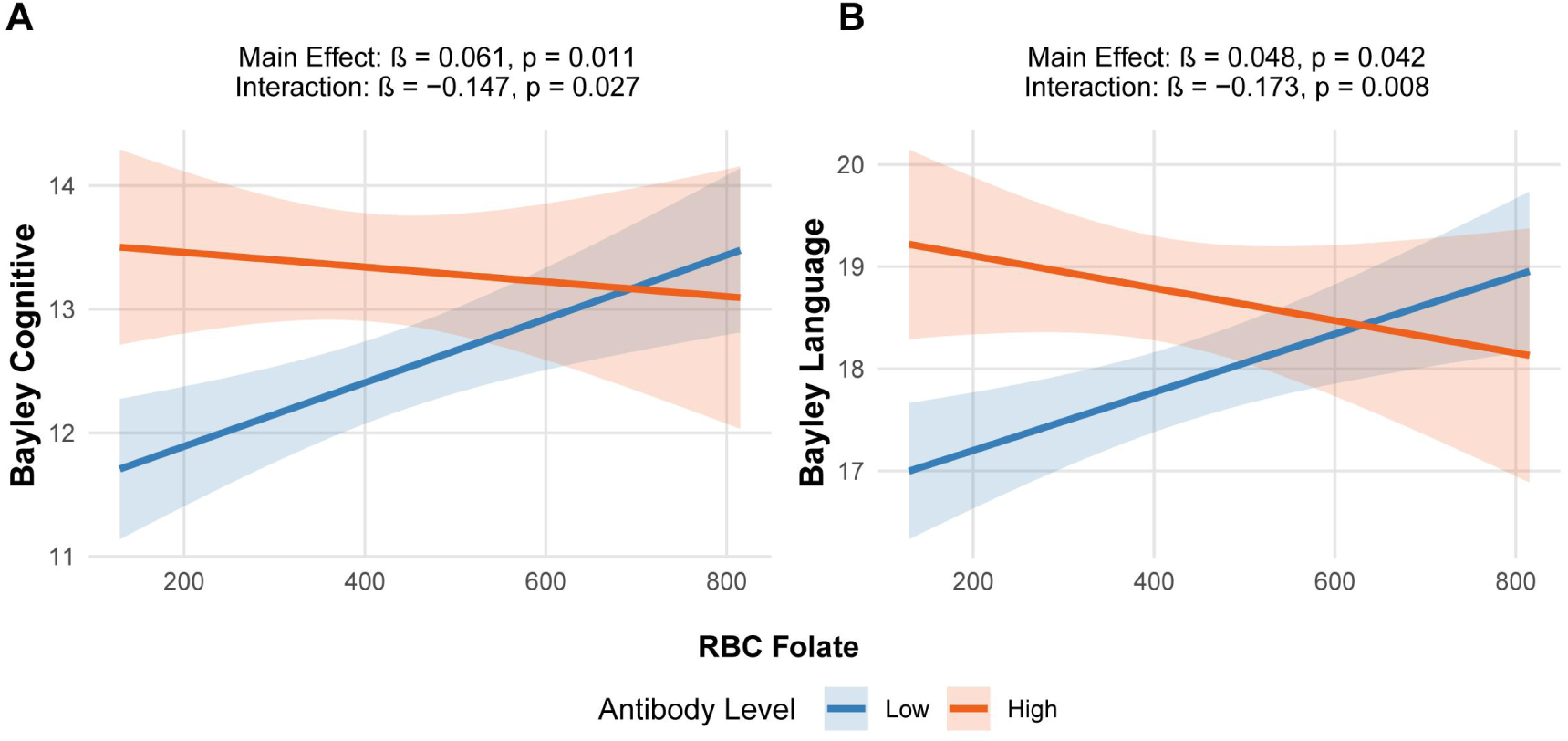
Interaction effect of high folate antibodies on the association between maternal folate and offspring Bayley scores at age 2. Participants were stratified into high and low antibody groups based on the 70th percentile (163.87 ng/mL) of folate antibody levels.

**Fig. 3.**
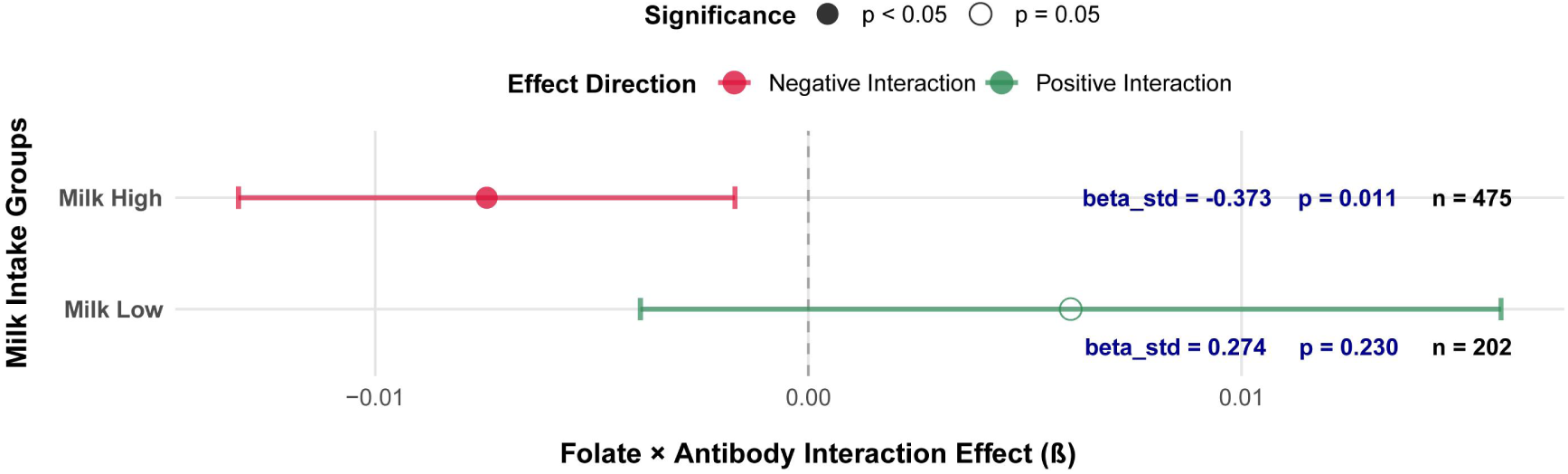
Milk Intake Specifically Enhance Antibody-Mediated Interference with Folate’s Cognitive Benefits. Forest plot displaying the folate*antibody interaction effects across milk intake groups. Points represent effect estimates (β) for the interaction term between folate levels and antibody status on cognitive development scores. Error bars indicate 95% confidence intervals.Points are color-coded to represent interaction effect direction (red: negative effects where antibodies attenuate folate benefits; green: positive effects) and shaped by statistical significance (solid: p < 0.05; hollow: p ≥ 0.05).

### Stratification by Maternal Milk Intake

The moderating effect of FRAA was specific to mothers with high milk intake. In this group, a significant negative interaction was present between RBC folate and FRAA status on cognitive scores (*β* = -0.373, *p* = 0.011). No such interaction was found in the low milk intake group (*β* = 0.274, *p* = 0.230).

### Genetic Architecture of FRAA

In a genome-wide association study, suggestive loci for FRAA concentrations were identified in genes involved in xenobiotic transport (SLC22A8) and immunogenetic regulation (the HLA region). Subsequent two-sample MR analysis found no evidence that genetically predicted milk intake causally influences FRAA levels in the general population. Detailed results are presented in the Supplementary Results.

## Discussion

Based on data from the SBC, which includes precise quantification of the active folate form (5-MTHF) by mass spectrometry, FRAA levels, and multi-time-point neurodevelopmental assessments in 3,006 mother-child pairs, this study confirmed the broad benefits of maternal RBC folate on offspring neurodevelopment and demonstrated their attenuation by FRAA. Furthermore, we found that this antibody-mediated attenuation was specifically amplified by higher maternal milk intake. These findings establish a new perspective for understanding the role of folate in prenatal neurodevelopment, highlighting maternal immune status as a key intrinsic factor influencing folate utilization efficiency. They indicate that assessing the neurodevelopmental benefits of folate requires consideration of the interplay between maternal immune status and nutrition, offering a novel viewpoint for prenatal nutritional interventions.

This study elucidates the relationship between maternal RBC folate and offspring executive function from both neuropsychological and neurodevelopmental perspectives. At the phenotypic level, association and enrichment analyses indicated a positive association of folate with neurodevelopment that was specific to the cognitive domain, particularly in executive function. At the brain level, fNIRS further revealed that higher maternal folate levels were associated with a steeper developmental increase in activation in the left middle frontal gyrus (channel 7), a key brain region supporting executive function, during a Go/No-Go task from ages 4 and 8 (*β* = 0.160, *p* = 0.014). This suggests that folate is associated not only with executive function performance but also with the developmental maturation of the brain regions that underpin it. Methodologically, a key strength of this study was the specific quantification of RBC 5-MTHF using LC-MS/MS. This approach provides dual analytical and biological advantages. Biologically, 5-MTHF is the primary circulating active form of folate, and its concentration in RBCs serves as a stable, integrated indicator of long-term folate bioavailability(15). Analytically, the specificity of LC-MS/MS for 5-MTHF excludes confounding by inactive folate metabolites, thereby providing reliable technical assurance for delineating the true association between bioactive folate status and higher-order cognitive functions(13). Thus, by leveraging a precise biomarker of bioactive folate with neuroimaging, this study provides multi-level evidence establishing a specific connection between maternal folate status and the development of the neural basis of executive function in offspring.

Maternal immune status, particularly the presence of FRAA, is a key intrinsic factor influencing the neuroprotective efficacy of folate. Current prenatal folate management primarily focuses on the dosage and timing of exogenous supplementation, while assessment of maternal immune status, especially FRAA screening, has not been incorporated into routine prenatal care systems, with related screening mostly confined to specific populations such as individuals with infertility(23). NNotably, our study in a large, prospective cohort representative of the general obstetric population identified that approximately 30% of pregnant women had FRAA levels above a threshold (70th percentile, 163.87 ng/mL) that significantly attenuated the positive associations between maternal RBC folate and offspring cognitive outcomes at age 2 (Bayley cognitive score: *β* = -0.147, *p* = 0.027; language scores: *β* = -0.173, *p* = 0.008). This demonstrates that clinically relevant FRAA levels are not restricted to specialized clinical groups but are found in a substantial portion (∼30%) of the general pregnant population in our cohort, where they can compromise the neurodevelopmental benefits of folate. Existing research suggests that FRAAs may hinder folate transport to the brain through mechanisms such as steric hindrance or receptor downregulation(24). When present in the mother, FRAAs can also block folate transfer to the fetus and disrupt its uptake into the developing brain(9), which aligns with clinical reports linking elevated gestational FRAA to adverse outcomes including neural tube defects(25), cerebral folate deficiency (11), and autism spectrum disorder(26). Our findings extend this evidence by indicating that FRAAs likely impair the promotion of offspring executive function and other higher cognitive domains by interfering with effective folate delivery to fetal brain tissue. Collectively, this evidence underscores the need to refine prenatal folate assessment. Moving from reliance on folate concentration alone toward a combined evaluation of folate levels and FRAA status could deepen our understanding of individual differences in folate utilization efficiency and their impact on offspring neurodevelopment.

The effect of dietary milk intake on neurodevelopment exhibits a distinct dependence on maternal immune status. Mothers with FRAA levels at or above the 70th percentile were categorized as “Immune-sensitive,” indicating a state of potential immune-mediated folate utilization impairment, whereas those with lower levels were considered “Immune-normal.” We found that higher milk intake specifically amplified the folate-disrupting effect of FRAA on child cognition, revealing a novel diet-immune interaction governing folate efficacy in neurodevelopment. Our study is among the first to show that milk, a common food, may exert opposing effects on depending on neurodevelopment depending on maternal immune status. The underlying mechanism may involve molecular mimicry: bovine milk contains a FBP structurally similar to human FRα(10). Clinical observations that milk-free diets reduce FRAA titers, which rise upon milk reintroduction(11), suggest that milk FBP could trigger or sustain this autoimmune response. This provides a plausible explanation for our stratified findings: in “immune-normal” mothers, milk likely offers nutritional benefits. In contrast, in “immune-sensitive” mothers, the same exposure may act as an environmental trigger, potentially exacerbating the antibody response and thereby impairing folate transport. The absence of a population-level causal effect of milk intake on FRAA levels in our MR analysis (Supplementary Results) further strengthens the interpretation derived from our stratified observational analysis, that the observed association likely represents a context-dependent effect occurring primarily in the susceptible "Immune-sensitive" subpopulation, rather than reflecting a ubiquitous causal relationship operating across the entire population. Our GWAS findings offer a potential biological explanation for these observations (Supplementary Results). The analysis indicates that genetic susceptibility loci for FRAA are enriched in the HLA region, involved in immune regulation, and in the SLC22A8 gene, implicated in xenobiotic transport. This suggests that an individual’s tendency to develop an FRAA response to dietary exposures such as milk may be influenced by genetic background: variation in HLA could affect immune recognition of milk proteins, while variation in transporters such as SLC22A8 might alter the internal exposure to relevant antigens. Together, these pathways may collectively contribute to a genetic predisposition for the “Immune-sensitive” phenotype, potentially explaining why only a subset of individuals carrying certain risk genotypes shows elevated FRAA and related folate disruption in the context of high milk intake. Therefore, adopting low-allergenic dairy alternatives may serve as a targeted intervention for Immune-sensitive individuals to reduce FRAA production and restore folate function, thereby shifting the paradigm from uniform dietary standards toward personalized recommendations.

### Limitations and Future Directions

A few limitations of our study should be noted. First, the functional mechanisms of folate antibodies require further validation through experimental models. Second, self-reported dietary and developmental data may be susceptible to recall and reporting biases. Third, residual confounding from unmeasured factors, such as additional micronutrient status, cannot be ruled out. Fourth, the operational definition of “Immune-sensitive” was based on an exploratory, data-driven threshold for FRAA levels. In the absence of a clinical cutoff, we systematically tested percentiles from the 50th to the 90th, selecting the threshold that showed the strongest statistical interaction with RBC folate on cognitive outcomes after Bonferroni correction. While this approach allowed us to identify a potentially meaningful subgroup, the threshold remains hypothesis-generating and requires validation with direct immunological assays and in independent cohorts..

## Conclusion

Our study demonstrates that FRAA significantly impair the neurodevelopmental benefits of maternal folate. This attenuation is moderated in a context-dependent manner by milk intake, being specifically amplified in “immune-sensitive” mothers. These findings highlight the need to incorporate FRAA assessment into prenatal care alongside conventional folate monitoring. Adopting such a personalized approach that accounts for maternal immune status and specific dietary factors such as milk intake will enable more effective strategies for optimizing offspring neurodevelopment.

## Data Availability

Access to the deidentified participant research data must be approved by the research ethics board on a case-by-case basis, please contact the corresponding author F.L. for assistance in data access request.

## Code Availability

Code for the replication of analyses conducted in the manuscript can be retrieved at https://github.com/Trancy961202/SBC_FRAA.

## Acknowledgements

This study was supported by grants from the National Natural Science Foundation of China (No.s 82430104, 82125032, 32441107 and 82272079), the China Brain Initiative Grant (STI2030-Major Projects 2021ZD0200800), the Science and Technology Commission of Shanghai Municipality (YDZX20253100003001, 23Y21900500, 23DZ2291100 and 2018SHZDZX01), the Shanghai Municipal Commission of Health and Family Planning (GWVI-11.1-34, 2020CXJQ01, 2018YJRC03), Innovative research team of high-level local universities in Shanghai (SHSMU-ZDCX20211100), Discipline Peak-Climbing Plan of Xinhua Hospital Affiliated to Shanghai Jiao Tong University School of Medicine(XKPF2024A50011), the Program of Shanghai Academic Research Leader (No. 23XD1423400), and the Shanghai Municipal Health Commission and Early Life PLan Program of Xinhua Hospital, Shanghai Jiao Tong University School of Medicine.

## Author contributions

Q-Y.L. and F. L. had full access to the data. F. L. and Q.L. conceptualized and designed the study. Q-Y.L., W.X., J.A., L.C., and C.H. curated and analyzed the data. F.L., Q.L., and Q.Z. interpreted the results. Q-Y.L., W.X. and Q.L. drafted the manuscript. All authors critically reviewed the manuscript. All authors had final responsibility for the decision to submit for publication.

