## Supplemental Material for "Rethinking Prenatal Folate Assessment: Maternal Folate Receptor Autoantibodies and Child Neurodevelopment"

Li, *et al.*

**Supplementary Methods**

**fNIRS Data Acquisition and Analysis**

fNIRS data were collected at the 4-year and 8-year follow-up visits during both resting-state and go/no-go tasks using the NirScan-900D system (Danyang Huichuang Medical Equipment Co., Ltd., Jiangsu, China). The device employed near-infrared light–emitting diodes (LEDs; 730 nm and 850 nm) as light sources and avalanche photodiodes as detectors. Data were sampled at 11 Hz. The system consisted of 24 light sources and 28 detectors, yielding 80 measurement channels with an average source–detector separation of 3 cm (range 2.7–3.3 cm).

*Resting-State Data*

Preprocessing of resting-state fNIRS data and functional connectivity analysis were performed using the Homer3 software package. Channels with poor signal quality were excluded based on amplitude (0.1–500), signal-to-noise ratio (SNR < 2), and source–detector separation (0–45 mm). Raw light intensity signals were converted to optical density (OD). Motion artifacts were detected using a 0.5 s detection window, 1.0 s time padding, a standard deviation threshold of 5.0, and an amplitude threshold of 0.5, and corrected using spline interpolation (parameter = 0.99). To reduce low-frequency drift and high-frequency noise, OD data were band-pass filtered (0.01–0.1 Hz). OD signals were then converted into concentration changes of oxygenated (HbO) and deoxygenated hemoglobin using the modified Beer–Lambert law, with partial pathlength factors set to 6.0 for each wavelength. A correlation-based signal improvement method was applied to further reduce motion-related noise. Functional connectivity was calculated by computing pairwise Pearson correlation coefficients between HbO signals across channels, generating a correlation matrix representing inter-channel resting-state connectivity.

*Go/No-Go Task Data*

Preprocessing of task-based data followed standard procedures. Raw optical intensity signals were converted to hemoglobin concentration changes using the modified Beer–Lambert law. Task blocks were segmented according to experimental markers, with Go and Go/No-Go conditions defined by block onset and offset times of equal duration. Data cleaning involved removal of NaN values, spike detection, identification of poor-quality channels through cardiac signal inspection and SNR estimation, and interpolation of bad channels using values from adjacent high-quality channels. Noise reduction included detrending, discrete wavelet transform for motion artifact correction, and principal component analysis (PCA) to remove global systemic interference. Signals were further band-pass filtered to attenuate physiological noise. Task-related activation was then estimated using a general linear model for go and go/no-go conditions.

**Genome-wide Association Study and Mendelian Randomization Methods**

*Genotype Data, Imputation, and Quality Control*

We conducted a genome-wide association study (GWAS) to identify genetic variants associated with FRAA concentrations among female participants in the Shanghai Birth Cohort (SBC). Genotyping was performed using the Illumina Global Screening Array–24 v3.0 (GSA-24v3-0-EA). Phasing and imputation were carried out through the Michigan Imputation Server using the 1000 Genomes Phase 3 v5 reference panel (GRCh37/hg19). Eagle v2.4 was used as the phasing engine, the allele frequency check was performed under East Asian mode, and variants with an imputation quality score (R²) below 0.3 were initially excluded.

Post-imputation quality control was performed using PLINK 1.9 following the Neale Lab imputation pipeline as described online. Variants with an INFO (R²) score below 0.8, a minor allele frequency less than 0.05, or a missing genotype rate exceeding 0.05 were removed. Variants deviating from Hardy–Weinberg equilibrium (p < 1×10⁻⁷) were also excluded to minimize genotyping errors. Duplicate and non-biallelic single-nucleotide polymorphisms (SNPs) were eliminated for downstream analyses.

Sample-level quality control included filtering individuals with missing genotype rates greater than 5%, identifying and removing heterozygosity outliers defined as ±3 standard deviations from the mean, and excluding related individuals closer than the third degree. Because all participants were Han Chinese females, ancestry-based filtering and sex-chromosome checks were not required. Only autosomal variants were retained for analysis. Principal components (PCs) were subsequently derived from linkage-disequilibrium–pruned autosomal SNPs to account for potential population stratification.

Following these quality control procedures, a total of 8.35 million high-quality genetic variants and 1,635 participants were retained for the final genome-wide association analysis. The genomic inflation factor (λ) was 1.01, indicating minimal bias from population stratification.

*Phenotype Definition and Covariates*

FRAA phenotypes were obtained from SBC, and normalized as continuous antibody_value measures. Covariates included age in years and the top 10 principal components (PC1–PC10) from PCA analysis. Because the dataset included only females, sex was not modeled as a covariate.

*Association Testing*

Association testing was performed using linear regression under an additive genetic model:

$$FAA level=\beta_{0}+\beta_{1}\left( SNP \right)+\beta_{2}\left( age \right)+\sum_{i=1}^{10} \beta_{PC_{i}}\left( {PC}_{i} \right)+\epsilon$$

Genome-wide significance was defined as P < 5 × 10⁻⁸ and suggestive significance as P < 1 × 10⁻⁵. A Manhattan plot and quantile–quantile plot were generated to visualize association signals. The GWAS summary statistics were shown in Supplementary Table 5 and used for downstream Mendelian Randomization (MR) analyses with UK Biobank milk intake.

*Mendelian Randomization Analysis*

To explore whether genetically predicted milk intake exerts a causal influence on circulating FRAA levels, a two-sample MR analysis was conducted. Summary-level data for milk intake from the UK Biobank (Neale Lab, v3) were used as the exposure dataset, and the Michigan-imputed FRAA GWAS served as the outcome dataset. Independent genetic instruments associated with milk intake at a significance threshold of P < 1 × 10⁻⁵ were selected after linkage disequilibrium clumping (r² < 0.1 within a ±10 mb window). Harmonization between exposure and outcome datasets was achieved by matching chromosome, position, reference, and alternate alleles to ensure alignment of effect directions.

MR analyses were performed using the inverse-variance weighted (IVW) approach under a fixed-effects model as the primary estimator. The MR-Egger regression method was applied to detect and account for potential directional pleiotropy, and the weighted median (WM) estimator was used to provide robustness to invalid instruments. Instrument strength was evaluated using the per-variant F-statistic, with values greater than 10 indicating adequate strength. Heterogeneity was assessed with Cochran’s Q statistic, and the I²_GX index was used as an approximate indicator of weak-instrument bias. All analyses were carried out in R version 4.3.3 using the MendelianRandomization package (version 0.10.0). Given that the exposure and outcome data were derived from different populations, which may introduce biases due to population-specific genetic architectures, we also attempted to implement the MR-EILLS method to account for potential cross-population heterogeneity using the MR-EILLS package (version 0.1.0) (Hou L,et al., 2025). However, this method failed to converge across all tested parameter configurations.

**Supplementary Results**

**GWAS Association Signals for FRAA**

We identified multiple suggestive loci associated with FRAA levels at the threshold of P < 1 × 10⁻⁵. The two most prominent signals were located on chromosome 11 (102.76 Mb, within the SLC22A8 gene region) and chromosome 6 (32.9 Mb, within the HLA region) (Supplementary Fig. 4-5). No variants reached the genome-wide significance threshold (P < 5 × 10⁻⁸). Lead variants and association statistics for all suggestive loci are provided in Supplementary Table 5.

**Mendelian Randomization Analysis of Milk Intake on FRAA**

After harmonization, seven independent SNPs were retained as valid instrumental variables for milk intake (Supplementary Table 6). All MR analyses yielded non-significant causal estimates for the effect of genetically proxied milk intake on FRAA levels (Supplementary Table 7). Specifically, the primary inverse-variance weighted (IVW) method, as well as the MR-Egger and weighted median estimators, showed no significant association. The MR-Egger regression indicated no evidence of directional pleiotropy, and Cochran‘s Q test revealed no significant heterogeneity. We additionally attempted to apply the MR-EILLS method to account for potential cross-population bias; however, this approach did not yield a stable solution.

**Supplementary Figures**

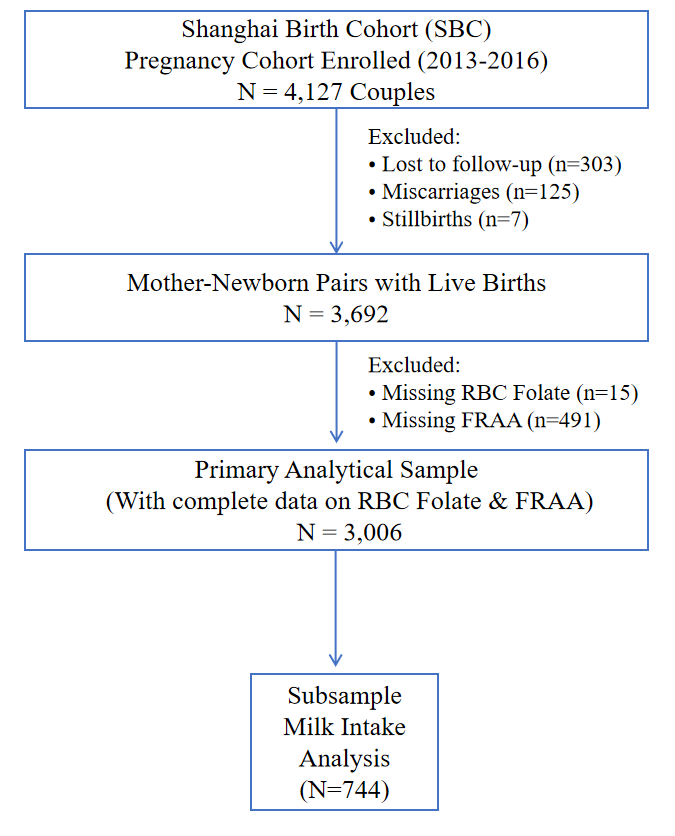

Supplementary Fig. 1. Study flow chart of the participant selection process in the Shanghai Birth Cohort (SBC).

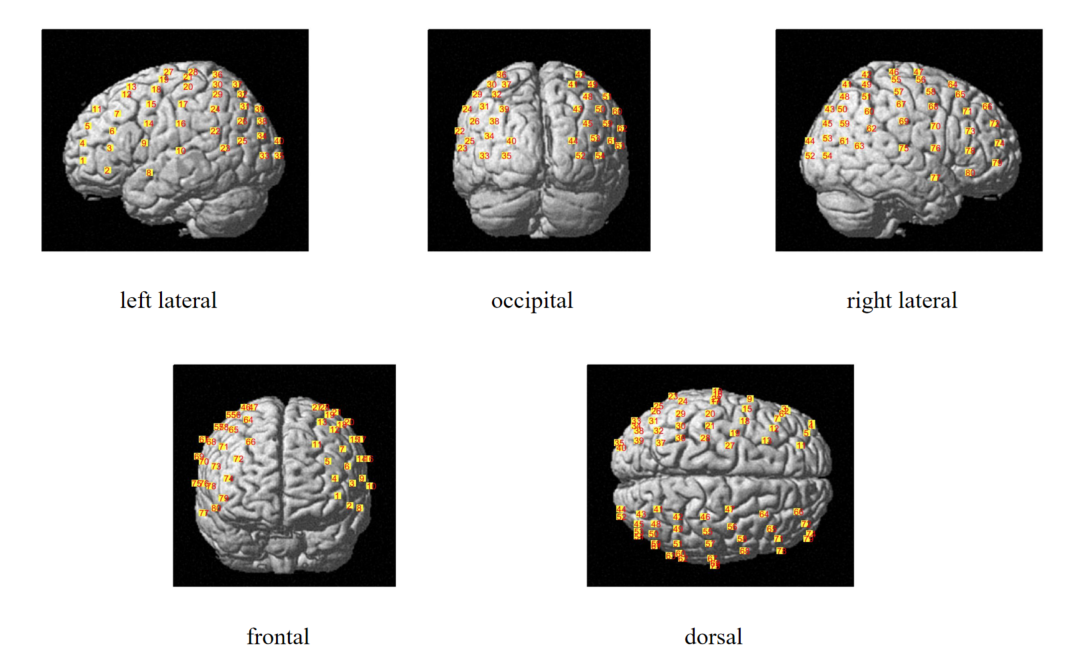

Supplementary Fig. 2. Layout and distribution of the 80-channel fNIRS montage.

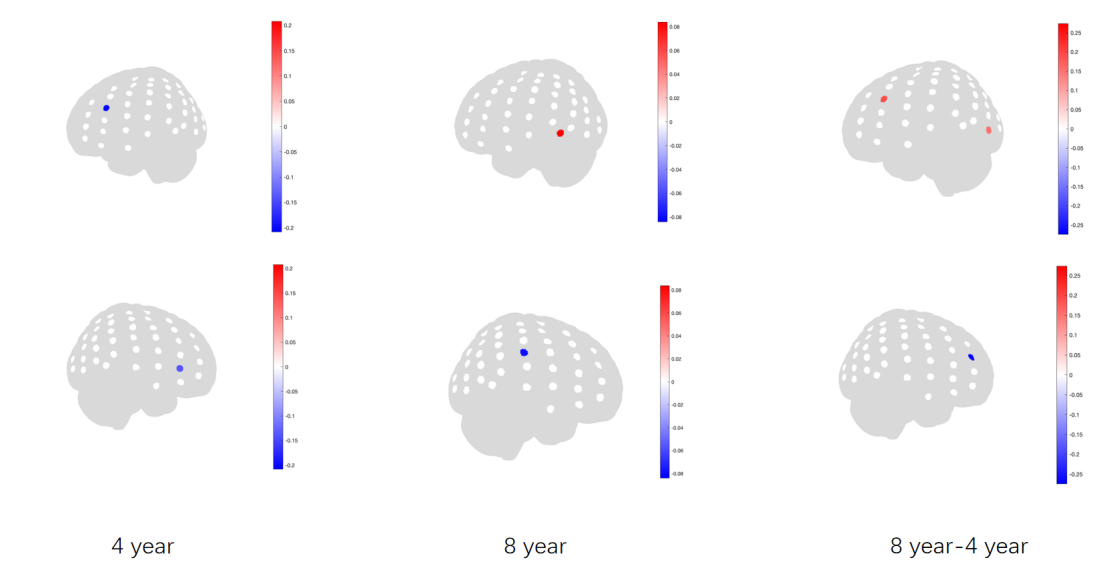

Supplementary Fig. 3. Significant correlation between folate and FNIRS.

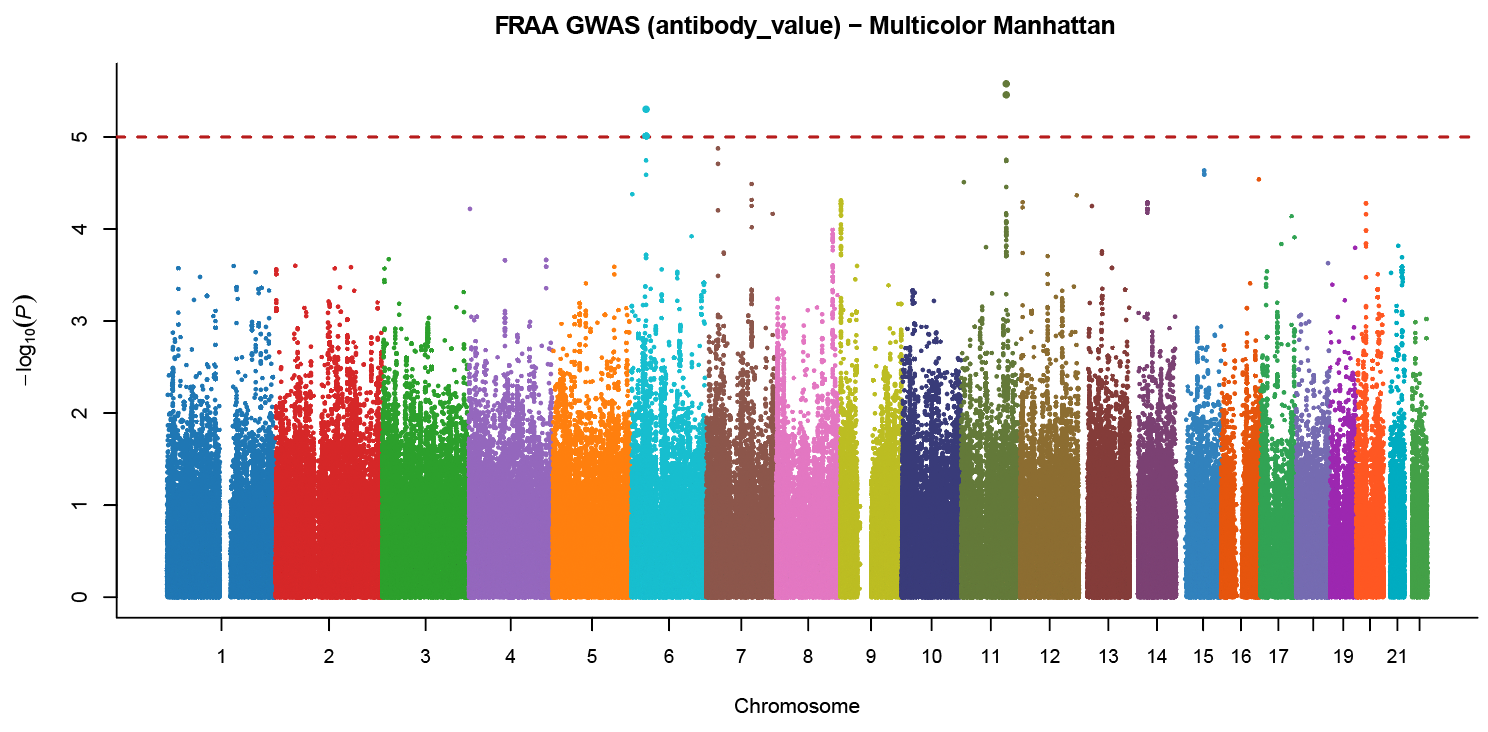

Supplementary Fig. 4. Manhattan plot for the folate receptor autoantibody (FRAA) GWAS.

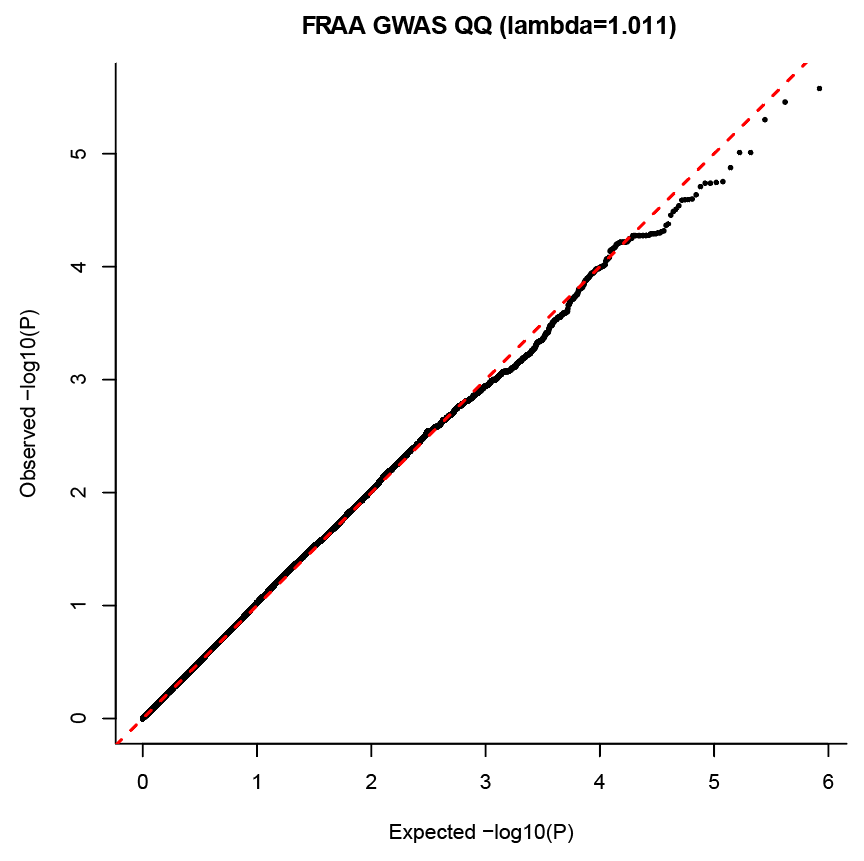

Supplementary Fig. 5. Q-Q plot for the folate receptor autoantibody (FRAA) GWAS.
